# Patterns of Polysubstance use in young Black and Latinx sexual minority men and transgender women in its association with sexual partnership factors: The PUSH study

**DOI:** 10.1101/2022.11.10.22282192

**Authors:** Renata Arrington-Sanders, Noya Galai, Oluwaseun Falade-Nwulia, Christopher Hammond, Andrea Wirtz, Christopher Beyrer, David Celentano

## Abstract

**Background:** Young Black and Latinx sexual minority men (SMM) and transgender women (TW) continue to suffer some of the highest burdens of HIV in the United States. Disparities are partly attributed to suboptimal uptake of HIV prevention and treatment services negatively impacted by substance use. Adult studies have demonstrated that polysubstance use increases HIV acquisition risk through increased sexual behaviors, however there are few studies that have examined polysubstance in this population and how polysubstance use might increase young SMM and TW’s risk for HIV.

**Methods:** Cross-sectional data from 466 young Black and Latinx SMM and TW living in four US high HIV-burden cities enrolled in the PUSH Study, a status neutral randomized control trial to increase uptake of pre-exposure prophylaxis and treatment adherence. Examined data for patterns of polysubstance use comparing age differences of use; and exploring associations between substance use and HIV risk behaviors focusing on three core partnership factors - inconsistent condom use, pressure to have anal sex without a condom, and older partner.

**Results:** Most participants described prior substance use with alcohol and cannabis being most common (76%, respectively) and 23% describing other describing prior alcohol use, 76% (n=353) described cannabis use, and 23% described other illicit drug use (including stimulants, cocaine, hallucinogens, sedatives, opioids, and inhalants). Polysubstance use was common with nearly half (47%) of participants reported alcohol and cannabis use, 20% reporting alcohol, cannabis, and one other illicit drug use, and 19% reporting alcohol or cannabis use plus one other illicit drug use. Polysubstance use was associated with greater adjusted odds of pressure to have anal sex without a condom, having an older partner (> 5 years older), and inconsistent condom use.

**Conclusions:** High levels of substance use, polysubstance use and strong associations with high-risk sexual practices and sexual partnerships that are known to be predictors of HIV acquisition or transmission among Black and Latinx sexual and gender minority youth, call for combination interventions that include substance use treatment alongside ARV-based prevention and treatment and partner-based interventions.

## 1.1. INTRODUCTION

Age and race-related disparities exist for sexual minority men (SMM) and transgender women (TW) in the US. In 2020, SMM accounted for 71% of new HIV infections with young Black and Latino SMM age 13-24 accounting for 54% and 27% of new infections in this age group. [1] Incidence rates are thought to be as high as 8.81-10.9 per 100 py in young Black SMM, 3.15 per 100 py in Latinx, and 3.75 per 100 py in mixed race youth. [2, 3] Transgender women accounted for 2% of new HIV diagnoses in 2020, with rates highest among Black and Latinx TW and 28% of new infections occurring among youth age 13 to 24. [1] Among Black and Latinx TW and nonbinary or gender queer youth, incidence rates may be around 2.91 per person years, slightly lower than some rates of Black SMM. [3] There has been a call to explore racial disparities, including partner factors associated with HIV incidence, including experiences of condomless anal intercourse, sexual coercion, substance and polysubstance use, that may predispose young SMM and TW to HIV.

Stimulant use is strongly associated with sexual HIV acquisition risk among SMM and risk is significantly increased with co-occurring binge drinking and cannabis use. [4, 5] Other work has identified methamphetamine use as a key predictor of HIV seroconversion among SMM. [6] In this study, men who use club drugs defined as (ketamine, ecstasy, GHB, cocaine, or methamphetamine) had a significantly higher number of missed PrEP doses compared to men who did not use club drugs, however, this work did not specifically explore the role of coexisting other illicit drug use in younger men. [6] Polysubstance use, defined as the use of more than one psychoactive substance of abuse, in particular, has been identified as a risk factor in HIV acquisition among adult samples of SMM and transgender women. [4, 7] Adult studies have demonstrated that polysubstance use increases HIV acquisition risk through increased sexual behaviors, such as group sex, [8] condomless anal sex, [9] and exchange sex. [10] However, little work has sought to understand the role of polysubstance use in sexual partnerships and whether the combination of polysubstance use might increase young SMM and TW’s risk for HIV. In a prior scoping review, young SMM were found to be more likely to engage in polysubstance use than older SMM in 3 of 5 studies that examined polysubstance use and sexual risk. [11] The 2 studies which found either no association or less polysubstance use in young SMM, used an imprecise measure of younger age (≤ 39 years) or focused only on Asian/Pacific Islander SMM. Lifetime substance use as high as 90% has been described among young transgender women, with some studies finding that over 50% of samples describe sexual intercourse under the influence of polysubstance use. [12-14]

Polysubstance use has been tied to sexual violence and condomless anal sex and can occur with and reinforce risk for HIV when it co-occurs with violence and sexual risk. [15-17] Both Black and Latinx SMM and TW experience age-discordant partnerships (e.g., partners >5 years older), sexual trauma, or abuse during adolescence, in the context of substance use which may further predispose youth to potential syndemics for HIV. [18-20] A syndemic is multiple co-occurring conditions (e.g., substance use, condomless sex) in the context of deleterious social and/or physical conditions (e.g., sexual violence, younger age/older partner) that mutually reinforce and increase vulnerability to an adverse condition (e.g., HIV). [21, 22] There are few studies that have explored patterns of polysubstance use among young SMM and TW over different age groups, and whether severity of use is associated HIV risk behaviors. Substance use among youth continues to be common with more youth engaging in polysubstance use. According to 2021 Monitoring the Future Survey [23] lifetime substance use among 10th and 12th graders in 2021 were respectively: 25.0 and 41.3% for illicit drugs, 34.7 and 54.1% for alcohol, 22.0 and 38.6% for cannabis, 5.2 and 4.9% for amphetamines, 3.5 and 7.1% for hallucinogens. Similarly, rates of use in the past year were: 18.7 and 32.0% for illicit drugs, 28.5 and 46.5% for alcohol, 17.3 and 30.5 for cannabis, 2.7 and 2.3% for amphetamines, 2.2 and 4.1% for hallucinogens. However, data presenting polysubstance use trends in this sample is limited.

Yet, no studies specifically examine these patterns across age for young Black and Latinx SMM and TW. The main objective of the current study is to characterize patterns of polysubstance use in young black and Latinx SMM and TW comparing age differences of use; and exploring associations between substance use and HIV risk behaviors focusing on three core partnership factors - inconsistent condom use, pressure to have anal sex without a condom, and older partner. We hypothesized that more severe substance use would be associated with greater sexual risk.

## 1.2. METHODS

### 1.2.1. Population & Recruitment

Participants were drawn from the PUSH Study of young Black and Latinx SMM and TW recruited from four cities in the US, (Baltimore MD, Washington DC, Philadelphia PA and Tampa/St. Petersburg FL) between August 2017 - June 2021. The study design is described in detail elsewhere. [24] Briefly, this was a mixed methods study with a qualitative, formative component, an initial cross-sectional survey and two embedded 18-months RCTs.

Recruitment has been described previously. [25] Briefly, participants were recruited using RDS with targeted seed selection, where seeds were identified through direct and indirect recruitment. Direct seed selection consisted of informational flyers directly given to a youth from study staff at a clinical site, community-based organization or in-person events. Indirect seed selection consisted of informational flyers posted at clinical sites that serve young SMM and TGW, offices of community-based organizations as well as a web-based approach with electronic ads placed on webpages, social and sexual networking sites frequented by young SMM (e.g., Jack’d, Black Gay Chat, Grindr). Eligibility for the initial survey was status-neutral and criteria included: (1) 15-24 years old; (2) reside in the four study sites; (3) assigned male sex at birth; (4) self-identified Black/African American or Hispanic/Latinx; and (5) report oral/anal sex with a man in the prior 12 months.

### 1.2.2. Measures/Assessments

All individuals meeting eligibility and provided voluntary study consent, completed a baseline survey and HIV testing. The baseline survey included questions about demographics (e.g., age, sexual orientation, gender identity, housing status), sexual behavior (e.g., number of partners, type of sex, etc.) drawn from prior measures [26], substance use, HIV history, engagement in preventive behavior (e.g., condom and PrEP use), disclosure of sexual orientation to provider, and city location of recruitment. Participants were offered a rapid oral HIV test (OraQuick Advance® or INSTI) at baseline, unless HIV status was known via medical record. The present analysis is restricted to participants who completed the baseline survey, were screened and found eligible to participate in one of the imbedded RCTs. Participants were excluded if they did not consent to the screening HIV test to determine their HIV status or if there was no verified test result on their medical records. Participants were paid $50 for completion of the baseline survey and the HIV test. The PUSH study protocol was approved by the Institutional Review Boards of all participating institutions including: The Johns Hopkins University in Baltimore, MD; Children’s National Medical Center and Whitman-Walker Health in Washington DC; Children’s Hospital of Philadelphia (CHOP), Philadelphia, PA; and Western IRB, for Metro Inclusive Health, St. Petersburg/Tampa, FL. Unique identifiers that included zip code was used to ensure participants were associated with only one geographical area.

### 1.2.3. Substance Use Measures

Substance use was measured using the CRAFFT a validated six-item screening tool designed to detect alcohol and drug use [27, 28] and the NIDA Modified (NM)-ASSIST. [29] Both instruments were used to account for the age range of the population. The CRAFFT is validated for use in adolescents [28] while the NM-ASSIST is validated for use in adolescents and young adults 18 years or older, although it has been used in adolescent populations. [30] CRAFFT score was categorized into – ‘0’(no dependence), and 1-2, and ≥3 based on prior literature in African American adolescents that suggest the use of a combination of cut-offs to demonstrate severity of substance dependence or problem consumption. [31] Participants answered the NM-ASSIST first for each substance to assess ever use followed by past 3-month substance use. For 3-month substance use, frequency (e.g., never, once or twice, monthly, weekly, daily or almost daily) was assessed for each substance. For this analysis, we examined lifetime and substance use in the past 3 months. The substances reported included tobacco, alcohol, cannabis, amphetamine type stimulants (speed, diet pills, ecstasy, etc.), cocaine, hallucinogens, sedatives, opioids, and inhalants. Drugs used with low frequency including amphetamine type stimulants (speed, diet pills, ecstasy, etc.), cocaine, hallucinogens, sedatives, opioids, and inhalants was further combined and categorized as other illicit drug use. To evaluate polysubstance use, we created a composite variable with categories based on combinations of substances reported to have been ever used (excluding tobacco): 1) both alcohol and cannabis; 2) alcohol and cannabis and at least one of the other illicit drugs; 3) alcohol or cannabis with one other illicit drugs; 4) no lifetime substance use. These categories were selected based on most frequently used substances reported by participants, which reflected prevalence of use in adolescents and young adults. [32]

### 1.2.4. Sexual Partnership Factors

Participants were asked to report sexual behavior (oral, anal, vaginal) in the last 3 months including sexual behavior with the last 4 partners. Participants were asked details about partners, including age, HIV status, and condom use for each partner and overall. Participants were also asked about pressure to engage in sex without a condom, experiences of coerced or forced sex, and history of transactional sex.

### 1.2.5. Variables and Exposures

Dependent variables represented several behavioral pathways for HIV acquisition or transmission and included inconsistent condom use, coerced condomless sex, and partner age discrepancy. Inconsistent condom use was based on the question “In the last 3 months, how often did you/your male partner wear a condom?” and was coded as 1 if the response was “Never” or “Half the time” and 0 if the response was “Always”. The outcome Pressured to have condomless sex was based on the question: “To what extent do you feel pressure from other people, including sex partners, to have anal sex without a condom?” It was coded as 0 if the response was “Not at all” and 1 otherwise (a little/somewhat/very much). Partner age discrepancy was defined as having a partner that was 5 years or older than the participant. The main exposure factor of interest was polysubstance use. Transactional sex was defined as affirmatively answering to the question, “Have you ever had sex with another male in exchange for money, place to stay or food?” Unstable housing defined as affirmative response to the following question, “In the past 12-months, have you been without a regular place to stay?”

### 1.2.6. Statistical Analysis

We first examined patterns of drug use using trend graphs over age and the distribution of specific drugs used within each category of polysubstance used. The association between polysubstance use and our defined dependent variables were examined with bivariate, and multivariate analyses. Bivariate analysis was performed using frequency tables and χ2 tests. Generalized linear models were fit to the binary outcomes with adjustment for clustering by geographic location. Multivariate logistic models with robust variance accounting for clustering within geographic site were developed for each outcome separately. Covariates known previously to be associated with sexual risk behaviors and potential confounders were included in statistical models including age, race/ethnicity, education, gender identity, sexual identity, and HIV status. The initial broad model included all sexual risk factors associated with the outcome with a p-value of <0.3, adjusted for clustering by city/geographic site; the second model examined substance use patterns, adjusted for clustering by city; and the third model examined patterns associated with substance use in the last 3 months. Model results are shown as odd ratios (OR), and associated p-values.

## 1.3. RESULTS

A total of 466 participants contributed to this analysis. The study population is described in Table 1. Participants came from all four cities. Mean age of participants was 21.3 years (2.45), most participants identified as African-American/Black (57%), with 15% (n=69) identifying as Latinx. Most participants self-identified as gay (n=277, 59%) or bisexual (n=115, 25%), completed high-school or higher education (78%), had a cell phone (92%), and health insurance (87%). Fewer participants (14%, n=67) identified as transgender or gender diverse. Twenty four percent of participants described unstable housing in the last year. Participants had an average of 23.0 total lifetime partners, with 50% reporting 10 or more lifetime partners.

**Table 1:** Description of study cohort: Participants demographic and socio-economic characteristics

| **Variable** | **N**<br>**(Total=466)** | **%**<br>**Total=100%** |
| --- | --- | --- |
| **City** |  |  |
| Baltimore | 154 | 33.05 |
| Philadelphia | 130 | 27.90 |
| Washington DC | 164 | 34.33 |
| Tampa/St Petersburg | 22 | 4.72 |
| **Age categories** |  |  |
| 15-17 | 47 | 10.09 |
| 18-19 | 104 | 22.32 |
| 20-21 | 110 | 23.61 |
| 22-23 | 123 | 26.39 |
| 24-25 | 82 | 17.60 |
| **Race (2 missing)** |  |  |
| African American/Black | 265 | 56.87 |
| Black other | 130 | 27.90 |
| Black Latino/Hispanic | 69 | 14.81 |
| **Education (1 missing)** |  |  |
| Less than high school | 102 | 21.89 |
| High school graduate | 196 | 42.06 |
| More than high school | 167 | 35.84 |
| **Health Insurance (3 missing)** |  |  |
| Parents insurance only | 198 | 42.49 |
| Own insurance only | 170 | 36.48 |
| Both parents & own insurance | 34 | 7.30 |
| No Insurance | 61 | 13.09 |
| **Being without a place to stay, last 12m (4 missing)** |  |  |
| No | 350 | 75.11 |
| Yes | 112 | 24.03 |
| **How well-off is your family? (2 missing)** |  |  |
| Well-off | 119 | 25.54 |
| Average | 217 | 46.57 |
| Not Well-off | 128 | 27.47 |
| **Gender Identity (1 missing)** |  |  |
| Male | 399 | 85.62 |
| Transgender woman | 32 | 6.87 |
| Other (Female/Non-Conforming/Questioning/Queer) | 35 | 7.51 |
| **Sexual Orientation (2 missing)** |  |  |
| Gay | 277 | 59.44 |
| Bisexual | 115 | 24.68 |
| Other (heterosexual, pansexual, questioning, queer) | 72 | 15.45 |
| **Disclosed sexual orientation to medical provider**<br>(1 missing) |  |  |
| No | 123 | 26.39 |
| Yes | 342 | 73.39 |
| **Dating status** (4 missing) |  |  |
| Not dating | 210 | 45.06 |
| Casual dating | 144 | 30.90 |
| Exclusive/cohabit/married | 108 | 23.18 |
| **Total lifetime male partners** |  |  |
| <=3 | 124 | 26.61 |
| 4-9 | 109 | 23.39 |
| 10-20 | 117 | 25.11 |
| >=21 | 116 | 24.89 |
| **History of transactional sex** (3 missing) |  |  |
| No | 360 | 77.25 |
| Yes | 103 | 22.10 |
| **HIV status**<sup>1</sup> |  |  |
| Positive | 140 | 24.46 |
| Negative | 326 | 63.95 |

<sup>1</sup>Note: HIV status was confirmed by a study HIV test for 88.4% of the participants; 26 of the youth with HIV and 28 of the at-risk participants HIV status was based on self-report.

The frequency of reported substance use is shown in Table 2. The minority(n=62, 14%) described no prior substance use. Nearly forty percent (38.3%) described tobacco use, 76% (n=354) described alcohol use, 76% (n=353) described cannabis use, and 23% described other illicit drug use. In Figure 1, frequency of using specific substances for each of the three groups defined by the combinations of reported use is reported for lifetime ever use (left panels) and the last three months use (right panels), with similar patterns emerging in both timeframes – nearly half (47%) of participants reporting alcohol and cannabis use, 20% of participants reporting alcohol, cannabis, and one other illicit drug use, and 19% reporting alcohol or cannabis use plus one other illicit drug use. Most participants who reported ever alcohol and cannabis use described using alcohol (98%) or cannabis (94%) in the last 3 months. Among participants that reported alcohol, cannabis and other substance use, the most common other substance used was cocaine (35%) followed by hallucinogens (30%). Age patterns emerged in the data (Figure 2), with younger adolescents (15-19 years old) describing more lifetime cannabis use than alcohol use, though with a notable inflection at age 21, in which young adult participants (20 years+) described more lifetime and past 3-month alcohol than cannabis. Over the age groups, amphetamine, cocaine, and sedative use increased in the higher age groups. Similar patterns were seen with reported use in overall and last 3 months use (Figure 2).

**Table 2:**
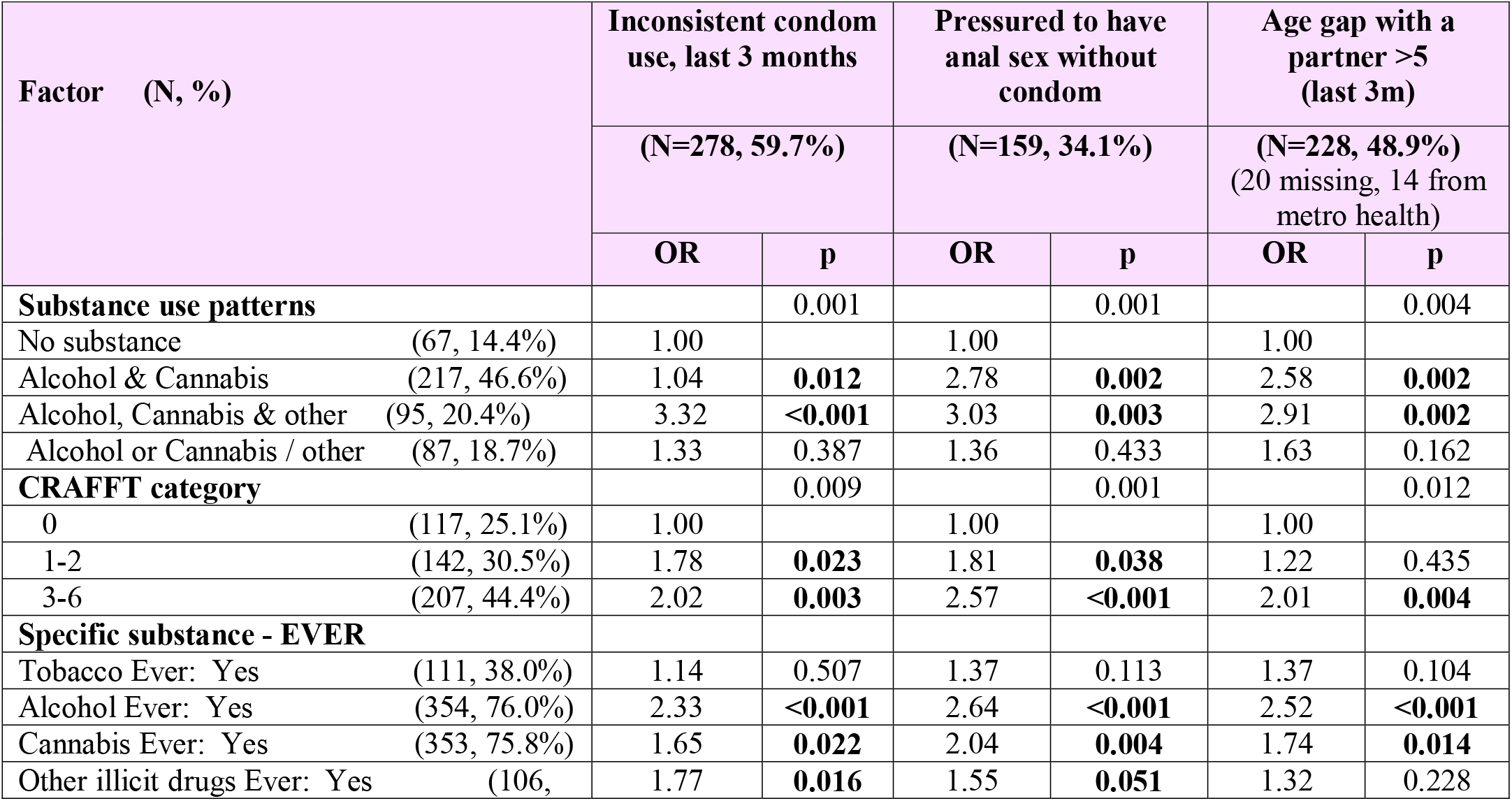
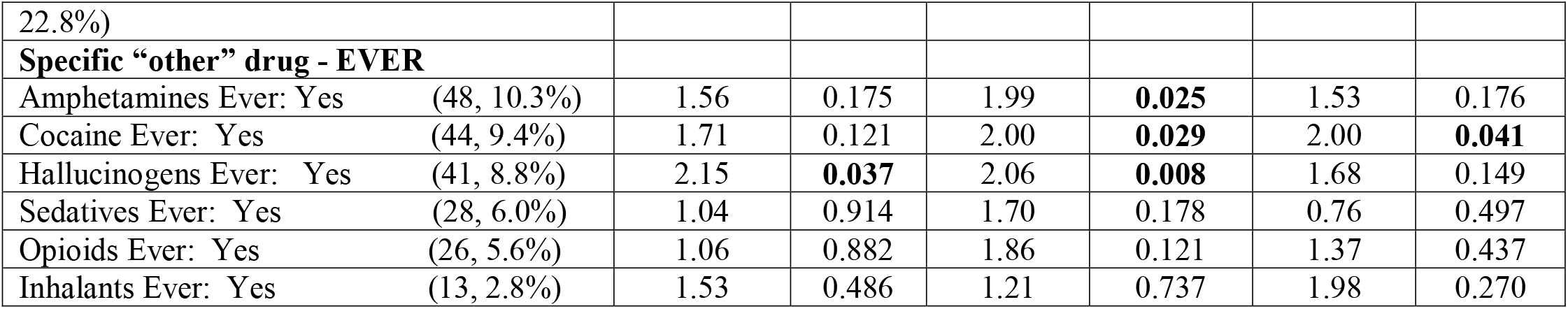
Bi-variate logistic model showing the association of Substance Use variables with Sexual Risk outcomes.

**Figure 1:**
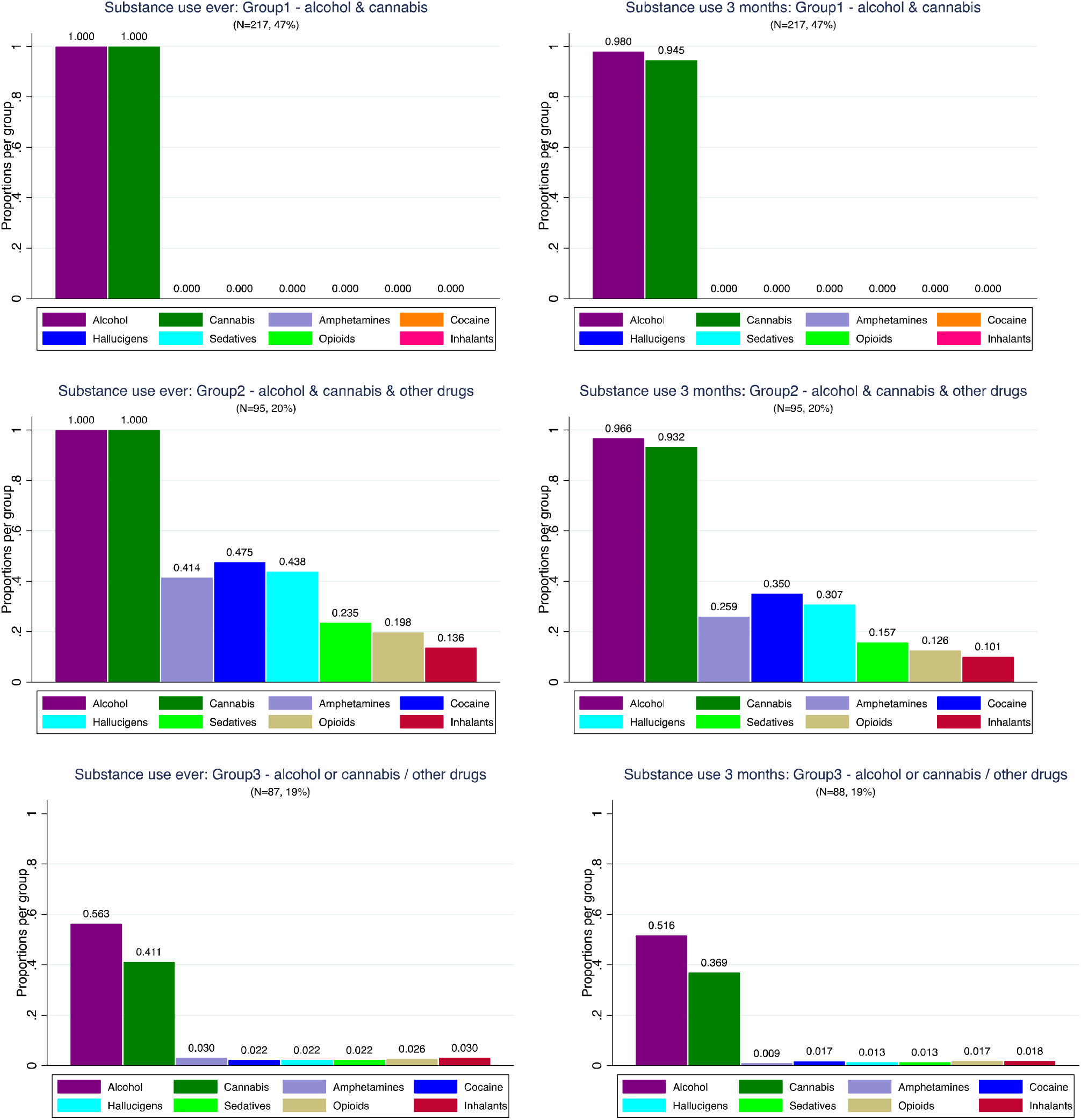
Substance use by groups of poly-drug combinations: Left panel – ever use; Right panel use in the last 3 months

**Figure 2:**
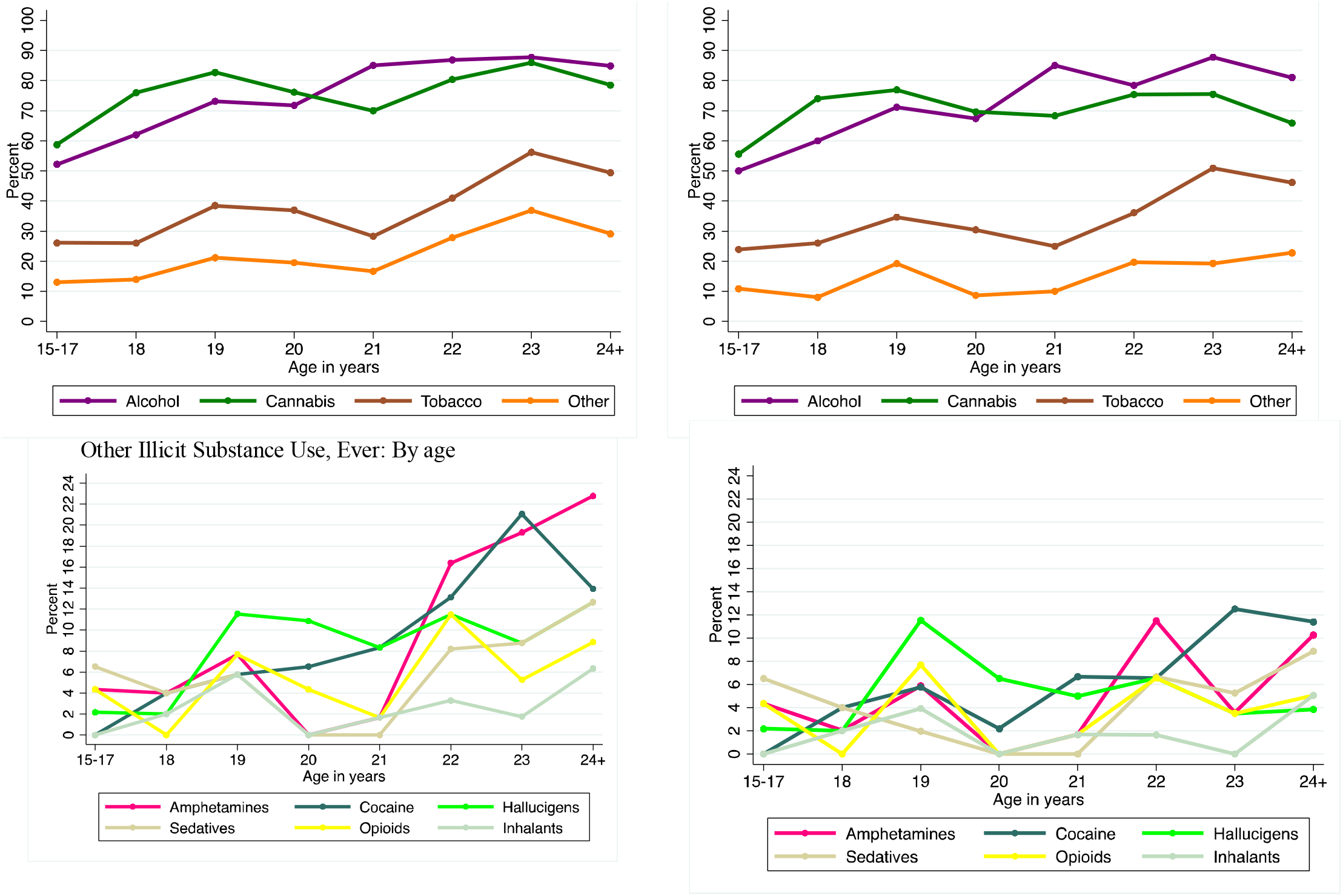
Substance use by age: PUSH (Other substances include: Amphetamines, Cocaine, Hallucinogens, Sedatives, Opioids, Inhalants)Other Illicit Substance Use, Ever: By age

The bi-variate association of substance use with sexual risk behaviors is shown in Table 2. Participants who described alcohol and cannabis had a 1.04 greater odds of inconsistent condom use, 2.78 greater odds of pressure to engage in anal sex without a condom, 2.58 greater odds of having a partner 5 years older in the last 3 months than non-substance using peers. This effect was magnified in participants who described alcohol, cannabis, and other substance use – where participants had a 3 times greater odds of inconsistent condom use, pressure to engage in anal sex without a condom, and have a partner 5 years older. Higher CRAFFT score was associated with inconsistent condom use, pressure to engage in anal sex without a condom, and a partner 5 years older. Patterns emerged around specific substance use with alcohol and cannabis independently associated with inconsistent condom use, pressure to engage in anal sex without a condom, and having a recent older partner. Amphetamine, cocaine, and hallucinogen use were associated with pressure to engage in anal sex without a condom, whereas cocaine use was also associated with having a recent older partner and hallucinogen use was associated with inconsistent condom use.

The association of substance use with the outcomes adjusting for participants’ characteristics and potential confounders is shown in Table 3. History of polysubstance use with alcohol and cannabis was associated with 2.34 greater adjusted odds of pressure to have anal sex without a condom and 1.68 greater adjusted odds of having an older partner. Reporting alcohol, cannabis and other substances was associated with 1.58 greater adjusted odds of inconsistent condom use, 2.35 greater adjusted odds of pressure, and 1.33 greater adjusted odds of having a partner 5 years older. Alcohol or cannabis use plus other substance use was not associated with outcomes in the final adjusted model. Independent relationships were seen for Latinx ethnicity, being out to provider, higher number of sexual partners and history of transactional sex.

**Table 3.** Adjusted association based on Multivariate logistic regression with clustering by city; Results are shown for factors that were significant for at least one outcome.

| Factor (N, %) | Inconsistent condom use, last 3 months |  | Pressured to have anal sex without condom |  | Age gap with a partner >5 (last 3m) |  |
| --- | --- | --- | --- | --- | --- | --- |
|  | aOR | p | aOR | p | aOR | p |
| **Substance use patterns (Ref: No substance use)** |  |  |  |  |  |  |
| Alcohol & Cannabis | 1.17 | 0.102 | 2.34 | **<0.001** | 1.68 | **<0.001** |
| Alcohol & Cannabis & other | 1.58 | **0.044** | 2.35 | **0.008** | 1.33 | **0.030** |
| Alcohol or Cannabis / other | 0.99 | 0.949 | 1.29 | 0.620 | 1.35 | 0.182 |
| CRAFFT (Ref: Score=0) |  |  |  |  |  |  |
| 1-2 | 1.26 | 0.421 | - | - | - | - |
| 3-6 | 1.18 | 0.481 | - | - | - | - |
| Race/Ethnicity (Ref: Black) |  |  |  |  |  |  |
| Latino | 2.43 | **0.003** | - | - | - | - |
| Disclosed sexual orientation to provider | 1.61 | **0.053** | - | - | 1.24 | **<0.001** |
| Number of lifetime male sexual partners (Ref: 1-3) |  |  |  |  |  |  |
| 4-9 | 1.38 | 0.206 | 1.80 | 0.124 | 2.51 | **0.006** |
| 10-20 | 2.78 | **<0.001** | 1.39 | **<0.001** | 3.10 | 0.059 |
| >=21 | 2.90 | **<0.001** | 1.72 | 0.105 | 9.27 | **<0.001** |
| Transactional sex | - | - | - | - | 1.24 | **<0.001** |

\*All models are adjusted for continuous age.

## 1.4. DISCUSSION

Participants in this sample of Black and Latinx SMM and TW described very high substance use overall and in the last 3 months. Participants who described severe or problem substance use, as marked by higher CRAFFT score, had greater odds of high-risk sexual behavior or partner experiences.

Polysubstance use was very common with 20% of participants reporting alcohol, cannabis, and one other illicit drug use, and 19% reporting alcohol or cannabis use plus one other illicit drug use. This rate is slightly lower to adult analyses that suggesting that non-medical prescription substance use with corresponding alcohol use may be as high as 27% lifetime use. [33] However, patterns of polysubstance use increased with older age with participants describing more illicit drug use with amphetamine, cocaine, and sedatives in the higher age groups in addition to alcohol and cannabis use. Polysubstance use was very common among participants who described both alcohol and cannabis use, with common substance including cocaine (35%) followed by hallucinogens (30%). These finding suggest that substance use in young Black SMM and TW may not be limited one substance, but multiple substances which may be predispose youth to different inherent risks for HIV acquisition.

Polysubstance use was independently associated with inconsistent condom use, pressure to engage in anal sex without a condom, and having a recent older partner. These relationships were independently seen among youth who reported a Latinx ethnicity, were out to their provider, reported higher number of sexual partners and history of transactional sex. In the context of substance use and intimate partner violence these findings suggest that more work is needed to explore and analyze power dynamics that may influence such relationships, and how experiencing multiple marginalized identities shapes one’s risk for substance use and associated adverse health outcomes.

Other data has demonstrated that the co-occurrence of high rates of condomless anal sex and substance use can predispose young SMM and TW to HIV. [12, 34] Experiences with polysubstance use and characteristics of sexual partners, for example older partnerships and pressure to engage in sex without a condom during substance use provide evidence that these youth may be particularly vulnerable to social contexts that predispose them to HIV. [35] Polysubstance use, combined with partner-specific factors like coerced sex and partners of discrepant age, may provide more opportunities for inconsistent condom use for youth. Youth who have engaged in transactional sex may inherently be particularly vulnerable to both intimate partner violence and polysubstance use, as such youth may use substances to cope or engage in transactional sex to support chronic use. The data in this sample also suggests that Latinx youth and youth with higher number of sex partners and more severe substance use may be particularly vulnerable to risk. This likely reflects a complicated relationship between more partners, which inherently increases one’s risk for more opportunities of inconsistent condom use, coerced sex, partners of discrepant age, transactional sex (which may be picked up by higher number of partners, even if not reported as such), and use of substances to cope or engage in transactional sex or to support chronic use. More research is needed to understand this relationship and whether polysubstance use in young SMM and TW is intermittent occurring predominantly only within sexual partnerships, chronic or as a means of coping.

Prior work has demonstrated that young Black sexual minority men may be more likely to engage in intermittent cannabis use than heavy use, but heavy use can be associated with lower awareness of HIV status. [36] Other work suggests that chronic substance use can impact cognitive functioning [37] and short-term memory. [38] Intermittent substance use may impact different mechanisms of decision making within partnerships and contribute to condomless anal sex and impaired communication around condoms. [39, 40] Combined with partner-specific factors, such as older partners and sexual coercion, younger SMM and TW may be particularly vulnerable to substance use at time of sex. Tailored interventions that address the different ways youth may be using multiple substances are needed to address the nuance of substance use within and outside of relationships. Such approaches would also require providers to address assumptions about the context of substance use in Black and Latinx SMM and TW and factors that contribute to sexual pressure or coercion within relationships, [41] including existing marginalized social identities (race, gender, sexuality, class, etc.) and having to navigate and experience larger macro systems of oppression (racism, heteronormativity, homophobia, etc.) that intersect to impact an individual’s health, relationships, and quality of life. [42]

Stimulant use, particularly cocaine and amphetamine use prior to or during sex has been associated with increased risk for HIV acquisition in SMM and TW. [43] Cocaine and amphetamine use has been reported predominantly during Chemsex for the pharmacological effects during sex (e.g., orgasm, enhanced pleasure). In this analysis, we found that 35% of polysubstance users also reported cocaine use. This rate is higher than other reports of stimulant use in young SMM and transgender women [15, 44] and may have significant implications in the participants in this sample. Participants who reported cocaine and amphetamine use had a greater odds of having experienced pressure to forgo the use of condoms during anal sex. Recent work by Xavier Hall suggests that young SMM and TW are likely to experience poly-victimization which can predict polysubstance use, particularly methamphetamine and cocaine use. [45] Partner-related coercive sex or pressure to engage in sex without a condom is a form of intimate partner violence experienced within relationships.

With prior work suggesting that younger SMM and TW may be at more risk for cocaine and amphetamine use prior to or during sex than older peers, [15, 46] interventions will need to be multi-factorial and address not only individual substance use, but also incorporate aspects victimization, violence and pressure that may occur within the sexual partnerships of young SMM and TW. [47] Continued research is needed to disaggregate components of HIV risk for young Black and Latinx youth – for example, how aspects of condomless sex, older partners and sexual violence may be inter-related to substance use. Intersectional screening tools are needed to alert providers to vulnerability, and training to be able to address the social context that youth experience which may include aspects of racism, homophobia, and heteronormativity that may increase substance use and impact power dynamics and vulnerability within sexual relationships.

### 1.4.1. Limitations

There are limitations of this work that should be noted. This data is cross-sectional limiting our ability to assess temporality or to determine prospectively how substance use changed over time or was modified by specific partnership factors within relationships. The samples are from geographically different, but mostly urban locations, limiting our ability to draw conclusions about similar youth in rural settings. More work is needed to delineate chronic polysubstance use from intermittent polysubstance use, because intermittent use during sexual partnerships may inherently present with different risks for HIV. We did not examine pre-exposure prophylaxis use as a dependent variable because we examined outcomes that would be relevant to both persons who were living with and at-risk for HIV. This limited our ability to draw conclusions around how substance use impacted protected and unprotected anal sex. Despite these limitations, this work suggests that in some samples of young Black and Latinx SMM and TW, there are very high levels of polysubstance use which may create a key vulnerability for HIV acquisition within partnerships.

### 1.4.2. Conclusions

Our data have demonstrated the high levels of substance use and polysubstance use marked by strong associations with high-risk sexual practices and characteristics of sexual partners that are known to be predictors of HIV acquisition or transmission among Black and Latinx sexual and gender minority youth. The relationships between substance use and inconsistent condom use, coerced condomless sex, and partner age discrepancies likely work through pharmacologic effects of these substances that are known to impair judgement, decision making, and negotiation alongside power imbalances within sexual partnerships. These findings highlight highly intervenable relationships that could be addressed through combination interventions that include substance use treatment alongside ARV-based prevention and treatment, partner-based interventions, IPV and safety planning services, and individual support to develop condom negotiation skills to address HIV disparities among Black and Latinx sexual and gender minority youth.

## Data Availability

All data produced in the present study are available upon reasonable request to the authors.

## List of abbreviations

SMM: sexual minority men
TW: transgender women
PrEP: Pre-exposure prophylaxis
HIV: human immunodeficiency virus
RDS: respondent driven sampling
Py: person years
OR: odds ratio

